# Social Support during pregnancy: A phenomenological exploration of young women’s experiences of support networks on pregnancy care and wellbeing in Soweto, South Africa

**DOI:** 10.1101/2022.04.03.22273162

**Authors:** Khuthala Mabetha, Larske Soepnel, Sonja Klingberg, Gugulethu Mabena, Molebogeng Motlhatlhedi, Shane A Norris, Catherine E Draper

## Abstract

Social support is deemed to have a crucial influence on maternal health and wellbeing during pregnancy. The objective of the study was to explore the experiences of pregnant young females and their receipt of social support in Soweto, South Africa. An interpretive phenomenological approach was employed to understand and interpret pregnant young women’s lived experiences of support networks on their pregnancy care and wellbeing. Data was collected conducting 18 indepth interviews with young pregnant women. Analysis of the data resulted in the development of two superordinate themes: (1) relationships during pregnancy and (2) network involvement. Each superordinate theme was linked to subthemes that helped explain whether young women had positive or negative experiences of social support during their pregnancy care, and their wellbeing. The sub-themes emanating from the superordinate theme ‘relationships during pregnancy’ were (a) behavioural response of partner following disclosure of pregnancy, (b) behavioural response of family following disclosure of pregnancy, and (c) sense of emotional security. Accompanying subthemes of the superordinate theme ‘network involvement’ were (a) emotional and instrumental support, and (b) information support. An interpretation of the young women’s experiences has revealed that young women’s satisfaction with existing support networks and involvement of the various social networks contributed greatly to the participants having a greater sense of potential parental efficacy and increased acceptance of their pregnancies. Pregnant women who receive sufficient social support from immediate networks have increased potential to embrace and give attention to pregnancy-related changes. This could, in turn, foster positive behavioural outcomes that encourage engaging in good pregnancy care practices and acceptance of motherhood.

Focusing on previously unexamined factors that could improve maternal health, such as social support, could improve maternal mortality rates and help achieve reproductive health accessibility universally.

## Introduction

Social support has been reported to play a crucial role in maternal health and wellbeing (1), influencing individuals’ ability to cope with events during specific periods (2). Social support is characterised by the manner in which connections that exist between people who have recurring interactions, fulfil the needs of individuals and this form of support can exist in an emotional, instrumental, affectionate or tangible form (3). The prenatal period is considered to be a delicate time particularly in view of the emotional, psychological and social wellbeing of expectant mothers (4). Pregnancy is generally viewed as a period that is coupled with various physiological and emotional changes, and these changes can have significant effects on both maternal and infant health outcomes (5-9). Close relations with other individuals play an integral role in influencing the quality of life of individuals which promotes positive wellbeing, subsequently resulting in expectant mothers perceiving pregnancy-related changes as less stressful (10). Thus, the care and support that pregnant woman receive contributes greatly to how they experience their pregnancy (11, 12).

A vast body of empirical evidence has been documented on the influence that social support has on the depressive symptoms that expectant women experience such as antenatal depression, postpartum depression, and subjective wellbeing amongst others (13-21). In particular, women who experience a lack of social contact or connection with others, and also perceive themselves to have low social support, have increased susceptibility to anxiety and antenatal depression, particularly if they have a strained relationship with their partner or family, which can subsequently affect various developmental outcomes of their babies (2, 22-30). Thus, social support may result in reduced risks of depression during pregnancy subsequently resulting in positive pregnancy and health outcomes (31).

Empirical evidence has shown that expectant mothers need support that involves undertaking tangible acts or activities, monitoring and care which promotes positive wellbeing and health for both mother and child (32). In addition, social support is deemed as important in strengthening positive health outcomes in families experiencing events that involve significant changes to their lifestyles such as the birth of a child or rearing of children (4, 33). Of note, significant differences have been observed with respect to the degree of social support received by adolescent mothers who are in the age groups 15 to 19 years and that of older mothers who are between the age groups 25 and above (23, 27), with results indicating that adolescent mothers received lower levels of social support than older mothers due to their weakened capability to create and sustain close connections and social ties with other individuals (23, 27). Despite the prevalence of adolescent pregnancy being largely documented in South African literature, stigmatization of adolescent pregnancy in the country remains rife (34). Empirical evidence has shown that stigmatization of adolescent pregnancy has the potential to weaken or compromise an adolescent’s social support systems, which influences the health of both the child and adolescent mother (35, 36).

Although there are various studies, particularly from high-income countries, that have examined social support during pregnancy (31, 37-39), most studies have been limited to adolescents or older mothers and most have examined social support from a quantitative perspective while there is a dearth of phenomenological inquiry that captures the lived experiences of social support through the in-depth narrative accounts of pregnant women. Additionally, studies that exist on social support have focused on partners’ role on the health of young mothers which has been limited to paternal involvement defined only through the state of being married or not married or the inclusion of the name of the father on the child’s birth certificate (6, 40-43). Moreover, most studies that focus on partner support have not been able to explain partner involvement and pregnancy outcomes as well as the type of support that partners offer, through more in-depth narrative accounts (6, 44, 45).

The potential role that families play in childbearing practices of young women merits further research given that the majority of South African adolescent and young women have been reported not to be in a marital union and are also resident in their families’ households during their pregnancy (46). Furthermore, several international studies have conducted research on social support initiatives that are specifically run by expectant mothers as well as professional initiatives during pregnancy (4, 47-52) leaving a gap regarding other forms of social support, such as family and peer support. The findings of these previous studies have shown that expectant females have obtained support from their peers and a therapeutic space in these support groups which has played a significant role in positive pregnancy outcomes and positive emotional wellbeing. In addition, pregnant women have received professional support through individualised education and supportive phone calls and other forms of support which are emotional, affirmational, informational and practical from both mother-to-mother support groups and healthcare providers, resulting in improved Quality of Life (QoL) during pregnancy (49). Also, receipt of social support has been mostly evident following the birth of a child (53). However, the need for social support is greater in the prenatal period.

It remains unclear what type of support is received by expectant mothers from low- and middle-income country settings, in particular young women, as there is limited literature on the social support patterns on the wellbeing of this specific age group and a lack of in-depth understanding of their relationships. Given this background, the study’s objective was to explore the lived experiences of young females’ receipt of social support and how it relates to their pregnancy care and wellbeing during the prenatal period, through a phenomenological approach, in the context of a South African township called Soweto.

## Materials and methods

### Study design

An interpretive phenomenological design was employed in this study. The rationale for employing this approach is that its philosophical and methodological foundations in phenomenology seek to offer in-depth insight into the experiences of individuals through describing, comprehending and interpreting participants’ experiences (54-56). Of note, individuals’ experiences are influenced by the world they live in, including certain determining factors in their lives which are often linked to social, cultural or political contexts (57). Thus, interpretive phenomenology was suitable in this context as it assisted in gaining insight into young women’s lived experience of receipt of social support regarding pregnancy care and wellbeing during pregnancy. Furthermore, this approach involves using semi-structured interviews and in-depth interviews and allows the researcher to search for meaning in the narrative accounts of participants (58).

This phenomenological view thus enables the researcher to try to make sense of the participant’s experience whilst gaining insight into the participant trying to make sense of what they have experienced (59). Moreover, the phenomenological method contributes significantly to the analysis of study participants’ narratives as it helps reveal the inner meaning or essence of a phenomenon (60). The phenomenological method employed here involves three central components: (1) bracketing, (2) analysis using imagination and instinct (3) describing (61). Bracketing implies that the researcher has preconceived notions, experiential knowledge or has certain expectations about the phenomenon under study but does not apply these elements during investigation (60).

Bracketing in this study was however applied by putting aside the researchers’ knowledge and beliefs about the phenomenon under study in order to accurately capture and describe the participants’ experiences. In terms of imagination and intuition, the researcher examines the phenomenon under study open-mindedly and the process of describing involves explaining or describing a phenomenon as accurately as possible without depending on any pre-given framework (62).

### Setting

The study took place at the Chris Hani Baragwanath Academic Hospital located in Soweto, Johannesburg, South Africa. It is nested in the Healthy Life Trajectories Initiative (HeLTI), and specifically the “*Bukhali”* randomised control trial that examines the effects of a complex intervention aimed at optimizing the health of young women preconception, during pregnancy, and postnatally (63). Soweto is a historically disadvantaged and underprivileged high-density peri-urban area that lies in the outskirts of the city of Johannesburg, with 1.3 million residing in the area (64). Although Soweto is characterized by diverse economic structures and activities, poverty related challenges remain rife in the area with food insecurity and unemployment being highly prevalent (65). The majority of the population residing in Soweto is deemed to have poor or limited access to appropriate healthcare services, particularly, youth-friendly services (66). Soweto is an area that can be deemed to be largely characterized by patriarchy in which gender inequality and harmful gendered stereotypes dominate childbearing decisions of young women (67-69). Additionally, the introduction of various health initiatives among young women during their reproductive years is pivotal given that 33% of young women will experience adolescent pregnancy and give birth to their first child before their 19^th^ birthday.

### Eligibility and participant recruitment

The study adopted a purposive sampling approach as it assisted in selecting participants who share similar characteristics and meet the selection criteria. Eighteen pregnant participants who were in the age groups 18 to 28 years, were recruited from the *Bukhali* trial and were interviewed. Participants assigned to the intervention arm received health literacy material pertaining to physical and mental health, micronutrient supplements, and free access to HIV and pregnancy testing. The intervention is delivered monthly by community health workers over a period of up to 18 months’ preconception, and continuing into pregnancy. Pregnancy specific material is provided to address healthy diet and healthy behaviours during pregnancy, and antenatal care activities that pertain to birth preparation and the child’s arrival (63). Participants in the control arm received information related to life skills and are also offered free HIV and pregnancy testing. Participants who had experienced a pregnancy loss were not recruited for this qualitative study. Due to the current blinding of the trial, combined results are reported for intervention and control participants.

### Data collection

Participants participated in individual in-depth interviews in the month of August 2021 at the research centre within the precinct of Chris Hani Baragwanath Academic Hospital in Soweto. Interview guide questions were developed by the study team and used for prompting where necessary. Interview questions focused on young women’s pregnancy experiences, support structures, health behaviours (antenatal care visits) and sources of information on pregnancy. Two local female interviewers fluent in both English and other South African vernacular languages conducted the interviews. Following COVID-19 safety protocols, the participants were interviewed in person in a comfortable and silent room that offered maximum privacy. An audio recorder was used and the interviews lasted between 45-60 minutes; notes were taken during each session in order to document key narratives and participants’ non-verbal cues. Prior to analysis, the recorded narratives of the participants were transcribed verbatim and interviews that were conducted in the participants’ native language were translated into English.

### Data analysis

Colaizzi’s seven-step method was used to analyze the data with the aid of MAXQDA software version 2020 (70) which assisted in recording and coding the interview transcripts. However, only six steps were employed in this study given that follow-up interviews did not take place in the seventh stage in order to validate our findings from the study participants. Colaizzi’s method is an inductive method used in phenomenology that assists in selecting prominent assertions, classifying and thinking logically of the meanings of the event or outcome being investigated as well as describing and depicting the participants’ experiences (71, 72). In the first step, we first checked the transcripts against the recordings and revisited the content so as to make sense of the lived experience of all the individual participants and to comprehend the feelings and thoughts of the young pregnant women. Next, we extracted significant phrases and statements from the transcripts that form a complete meaning of each participant’s experience. This was done by further re-reading each transcript in order to point out prominent assertions from each interview transcript. We then wrote separate assertions for each individual. In the third step, we formulated more general meanings from each significant statement on the text. We then coded and categorized these meanings from the various transcripts and checked for consistency of the meanings across all transcripts. In the fourth step, after acquiring meanings emerging from the prominent assertions, we then arranged meanings into thematic clusters. Accompanying subthemes are shown in Table 1.

**Table 1.** Themes focusing on young women’s experiences of social support on pregnancy care and wellbeing.

| Superordinate theme | Sub-themes |
| --- | --- |
| 1. Relationships during pregnancy | 1.1. Behavioural response of partner following disclosure of pregnancy<br>1.2. Behavioural response of family following disclosure of pregnancy<br>1.3. Sense of emotional security |
| 2. Network Involvement | 2.1. Emotional and Instrumental support<br>2.2. Information support (medical and cultural information support) |

Two superordinate themes were developed from the cluster of themes that adequately explained young women’s experiences of social support on pregnancy care and wellbeing. Superordinate themes and sub-themes were developed by mapping interrelationships, connections and patterns across the narratives through engaging with the recordings and revisiting each transcript several times. Several phrases were coded, and important phrases were identified resulting in the generation of the superordinate themes and sub-themes. In addition, for each superordinate theme, specific extracts for that theme were selected from different participants and placed under each relevant theme. This assisted us in achieving internal consistency and relative broadness of each superordinate theme. In the fifth step of analysis, we integrated all emerging themes into descriptions of the event or outcome being examined. This was done by bringing together all the thematic clusters, superordinate themes and generated meanings to create a structure that expressed the meaning in each theme (Table 1).

## Ethical Statement

Approval to conduct the study was granted by the Human Research Ethics Committee (Medical) based at the University of the Witwatersrand (M190449). Participants were requested to sign an informed consent form which stipulated the aims of the study, guaranteed confidentiality and indicated that the study is voluntary. The informed consent form also provided information on who would have access to the participants’ information, indicated that the study was voluntary, and provided information on the storage of data, recording of data and the dissemination of the findings. All participants who were willing to participate voluntarily and fully acknowledged that they understood the purpose of the research written permission to be interviewed, and for the narratives to be recorded.

## Results

All 18 participants were between the age groups 18 to 28 years. Overall, three participants had a primary school qualification, ten had a secondary school qualification, two had a tertiary qualification and three participants did not complete their primary education. Only six participants were enrolled in a higher education institution. Almost all females (16) indicated being in a relationship when the interview took place with only four of the participants living or cohabiting with their partners and 14 living with their families. Nine young women reported that they had given birth before, with the current pregnancy being their second pregnancy. Sociodemographic details are provided for each participant in Table 2.

**Table 2.** Demographic Profile of the participants.

| **Participant code** | **Current age** | **Highest level of education attained** | **Currently enrolled in Higher Education Institute** | **Employment status** | **Lives with** | **Relationship status** | **Number of pregnancies** |
| --- | --- | --- | --- | --- | --- | --- | --- |
| Participant 1 | 21-25 | Secondary school qualification | Yes | Unknown | Parents and siblings | In a relationship, not living together | 1 |
| Participant 2 | 21-25 | Secondary school qualification | No | Unemployed | Partner | In a relationship, and living together | 1 |
| Participant 3 | 16-20 | Secondary school qualification | No | Unemployed | Parent and siblings | Single | 1 |
| Participant 4 | 16-20 | Primary school qualification | No | Unknown | Parent, relative and siblings | In a relationship, not living together | 2 |
| Participant 5 | 16-20 | Primary school qualification | No | Unemployed | Grandparent and relative | In a relationship, not living together | 1 |
| Participant 6 | 21-25 | Primary school qualification | No | Employed | Sibling | In a relationship, not living together | 1 |
| Participant 7 | 21-25 | Tertiary qualification | Yes | Unemployed | Parent | In a relationship, not living together | 2 |
| Participant 8 | 21-25 | Secondary school qualification | No | Unemployed | Partner | In a relationship and living together | 2 |
| Participant 9 | 16-20 | Some primary school | No | Unemployed | Parent and sibling | In a relationship, not living together | 2 |
| Participant 10 | 21-25 | Secondary school qualification | No | Unemployed | Parent and siblings | In a relationship, not living together | 2 |
| Participant 11 | 21-25 | Secondary school qualification | No | Unemployed | Child | Single | 2 |
| Participant 12 | 21-25 | Tertiary qualification | Yes | Employed | Partner | In a relationship and living together | 2 |
| Participant 13 | 21-25 | Some primary school | No | Unknown | Partner | In a relationship, and living together | 1 |
| Participant 14 | 21-25 | Secondary school qualification | Yes | Employed | Parent | In a relationship, not living together | 1 |
| Participant 15 | 21-25 | Some primary education | No | Unknown | Parent and relative | In a relationship, not living together | 2 |
| Participant 16 | 16-20 | Secondary school qualification | Yes | Unknown | Parents and relatives | In a relationship, not living together | 1 |
| Participant 17 | 16-20 | Secondary school qualification | Yes | Unknown | Grandparents | In a relationship, not living together | 1 |
| Participant 18 | 26-30 | Secondary school qualification | No | Unemployed | Child | In a relationship, not living together | 2 |

### 1. Relationships during pregnancy

#### 1.1. Behavioural response of partner following disclosure of pregnancy

The narratives of the participants indicated that most of the participants’ pregnancies were unplanned. Despite this, most participants reported feeling happy about their pregnancies. Participants described feeling surprised and shocked when they learned about their pregnancy, and their subsequent meaning-making about their pregnancies tended to reflect overall relationship dynamics. The young women were worried about disclosing the pregnancies to their partners and feared getting negative reactions. However, many of the young women reported that their partners responded with great happiness and joy to the news of their pregnancy and their relationships with their partners remained stable. Thus, the partners’ behavioural response made the young women feel supported and safe in their relationships, fostering feelings of happiness and acceptance of the pregnancy and translating into positive wellbeing for the participants.

> “*Things are fine between me and my partner, normal as before. Since he accepted, he feels like he is okay, because he is also very supportive.” (*Participant 1, 21-25 years old).
>
> “*He is also excited because it’s what he’s been hoping for and to him it’s the first child We’re all okay, and also we stuck together, he’s supportive. We stay together, we do all things together, we’re not apart; only if he’s at work. Like we’re all on the same page*.” (Participant 2, 21-25 years old).

One of the young women’s narratives showed that her partner’s family had wanted her to fall pregnant which led to plans of marriage, although the young woman had not actually married her partner when the interview took place.

> “*The pregnancy was something that they wanted to plan, yeah, because the guy came to our home and took out some, it’s not lobola [customary payment for marriage], but he was just introducing himself and his family*” (Participant 6, 21-25 years old).

However, contrary to these findings, other participants indicated that their relationships with their partners changed following the disclosure of their pregnancy, as their partners’ reactions changed how the women viewed the relationship as opposed to how they viewed the pregnancy. Partners distanced themselves from the young women resulting in a dissolution of the relationship. Such circumstances resulted in the young women having negative wellbeing and not accepting their pregnancies.

> “*My relationship with my partner was fine, until I was pregnant. I told him about the pregnancy and he did not want to be involved, and I just cancelled him out of my life*.” (Participant 3, 16-20 years old).
>
> “*I told the child’s father that I am pregnant then he decided that we should stop seeing each other. He said to me in March, he does not want this child, he won’t be able to help me so it’s best that I get an abortion, so I asked him that when I get an abortion, am I supposed to do it alone, because after that you have to get me cleansed, and then he said to me that I will find my own way, he is not interested, I must just leave him alone*.” (Participant 4, 16-20 years old).

Another distinctive feature emerging in one of the narratives showed that although the young woman was receiving instrumental support from her partner, she had previously thought of terminating the pregnancy given the change in her partner’s behaviour towards her following her disclosure of the pregnancy to him.

> “*After I told him I was pregnant in April he started behaving strangely. He wasn’t warm*.” (Participant 13, 21-25 years old).

#### 1.2. Behavioural response of family following disclosure of pregnancy

Most participants reported that their families were shocked at the news of the pregnancy while some reported that their families were highly disappointed with them as they had wished that the young women could have focused on advancing their education or career. Despite this circumstance, the relationships between the young women and their family members remained stable and most were resident in the same households as their families and indicated feeling supported by their families and also had a sense of closeness which contributed positively to their pregnancy care and wellbeing. Most participants could confide and seek guidance from their families and this played a significant role in their pregnancy care and wellbeing:

> “*I thought that my parent was going to be angry or chase me away or something, but then my parent sat down and spoke to me and told me to keep it, because my parent has never made anyone do it, my parent is the first born so why does my parent have to judge me, life will go on and we will be alright*.” (Participant 4, 16-20 years old).
>
> “*My parents asked me that they heard my grandparent telling them that I am pregnant and I said yes I am pregnant. My parent said okay, there is nothing I can do since you are already pregnant. I will not tell you to abort, when you are pregnant, you are pregnant. A baby doesn’t have to be aborted and then my parents said that they will support me where they are able to support me, and that I will understand, and I told my parent that I don’t have a problem as long as you are going to support me, which is the only thing that I need, and that was it.”* (Participant 8, 21-25 years old).
>
> “*My relatives said whatever decision I take they will support me, whether I keep the baby or not*.” (Participant 16, 16-20 years old).

Contrary to these findings, the narrative of one participant showed that her relationship with her family changed drastically following disclosure of her pregnancy, resulting in a dismantled relationship and poor communication, and the participant experiencing negative emotions.

> “*My parent she was also disappointed, very disappointed I won’t lie but not as much as my other parent, my parent was really like shattered, like yoh in so much disbelief, my parent didn’t believe. It was hectic, hectic to a point where my parent cut me off financially, uh nobody spoke to me until now that I’m 7*… *like last month when I was 7 months; so nobody spoke to me basically for like 4 or 5 or 6*… *ja 3 months*.” (Participant 17, 16-20 years old).

#### 1.3. Sense of emotional security

The stories of the respondents indicated that they felt a sense of emotional security around their pregnancies. An interpretation of their narratives is that this sense of security and positive wellbeing stems from the fact that most participants have good social connections with family members and sexual partners. The presence of families and partners in the participants’ lives during their pregnancy has led into increased feelings of comfort. joy, satisfaction and positive self-awareness. This support and close relationships have also helped them to bond with their unborn babies and to have increased pregnancy acceptance and positive wellbeing.

> “*I’m happy, ja; like there’s nothing that I can think about because a baby is a gift and I had to accept that it’s mine. I have a life inside of me I must take care of myself*” (Participant 7, 21-25 years old).
>
> “*I feel great because like I’m carrying a fully-fledged human being, one that breathes and is alive, ja*” (Participant 5, 16-20 years).

### 2. Network Involvement

#### 2.1. Emotional and instrumental support

An exploration of the participants’ narratives showed that both partners and families of the pregnant young women are a key pillar in supporting the young women during their pregnancy and helping them navigate through challenges associated with pregnancy. An interpretation of their narratives shows that although the participants received financial support from their families and partners, participants mostly received emotional support which contributed significantly to their emotional wellbeing. The narratives also showed that some participants received most support from their partners, some from their families and some had an equal balance of support from both networks. In addition, partner and family engagements included provision of funds to meet healthcare needs, food security, and provision of a conducive living environment. These narratives suggest that partners and families demonstrated increased understanding and concern for maternal care and wellbeing of the young women during their pregnancies, which contributed immensely to positive pregnancy-related mental changes, being happy and accepting the pregnancy. This could potentially contribute to engagement in healthy behaviours, which will positively contribute to a successful delivery of the baby and positive post-partum care.

> “*The support I get from my parent, it is better. Makes everything easier. My parent cooks for me and makes sure I eat healthy*.” (Participant 10, 21-25 years old).
>
> “*My partner supports me, he’s there emotionally, physically, financially he supports me with everything that I need, he is there, we talk, his there, he is open, yeah. I am also under his medical aid.”* (Participant 6, 21-25 years old).
>
> “*He is just strong at being a very supportive partner. In terms of my education and job hunting and he has been watching my diet since pregnancy. He is my dietician on that one, emotional support he is there*.” (Participant 12, 21-25 years old).

Although Participant 12’s partner plays a key role in providing her with support in terms of her furthering her education, looking for employment and ensuring that she participates in healthy eating behaviours and providing for her financially, the participant indicated that her partner is not emotionally invested in the “physical pregnancy”.

> “*He doesn’t want to have those intimate experiences or moments with the baby. Because sometimes okay, the first time I experienced the baby kicking, I asked him to come and feel and he was just okay. He doesn’t ask much so I decided I’m no longer inviting him. He does ask on when last I felt the baby kicking and I would tell him that I felt it earlier on*.
>
> *He would then say I didn’t tell him but to me it would make no difference because he just touches my stomach and say okay!*”. (Participant 12).

In addition to receiving both instrumental and emotional care from their family members and sexual partners, two participants indicated also receiving support from the families of their sexual partners. Partners’ families showed concern, acceptance, affection and warmth to the participants and also provided them with emotional support which upon further interpretation suggests that they felt a sense of responsibility and obligation towards the unborn child.

> “*My partner, my family and partner’s family support me emotionally. When I have something that bothers me from home or with my friends, I speak to him, and he is able to support me. Makes me feel happy cause it is rare to find someone who is there with you and support you, especially during tough times. Yes, I am able to talk to them.”* (Participant 1, 21-25 years old).
>
> “*I think they are giving me love and they are trying to make me feel comfortable with the pregnancy, because at first I was like no, my age and a second child, I won’t be able to, and then my parent, my partner’s parent, they are there for me, that it’s something that happens, that you want to study further, it’s not something that will stop, like they give me, they encourage me you see, we are there, we will stay with the child, you can carry on with what you want to do and whatnot and whatnot, you see, so yeah*.” (Participant 10, 21-25 years old).

Despite these two narratives, an account of one participant showed that she has never had a meaningful connection with the father in either of her two pregnancies, nor received any instrumental help from him, his family or her own family. Such adversities have affected her emotional wellbeing resulting in her feeling less happy about her pregnancy.

> “*I’ve been having fights with the baby’s father, so I have lost interest in the pregnancy. Another thing is the lack of support from his family and mine. I sometimes feel depressed like sad, I lost hope and I even regret why I fell pregnant.” (*Participant 11, 21-25 years old).

#### 2.2. Information support

The majority of the participants reported receiving peer support through friends who provided support to the participants through sharing knowledge based on their own previous experiences, emotional support, social interaction or practical help and information on pregnancy care and pregnancy education from trial staff (details about intervention and control arms removed from quotes to protect blinding). An interpretation of the narratives brings forth the argument that the information support that the young women have received from the trial and health services has contributed greatly to them having positive feelings about their pregnancies, feeling less isolated and having improved emotional wellbeing; this may subsequently lead to increased parenting capabilities.

> “*I got most of the information from [the trial]. That is the one that made me understand most about the baby. That when the baby gets to certain months, they can listen to sounds, they can bond with you, the hair grows.”* (Participant 3, 16-20 years old).
>
> “*The information that I have is about the growth of the baby, the change of the woman’s baby, I got it from the clinic, obviously the card that I have, and then how the belly grows, I have got a book from [the trial], we went through a lot of pages, it shows which month the baby grows like this.”* (Participant 6, 21-25 years old).
>
> “*I got support from [the trial] and the clinic. On how to raise the child and having to breastfeed the baby up until six months for them to be healthy. Like how I am supposed to eat, I must avoid stress most of the time*.” (Participant 14, 21-25 years old).

Other participants reported that they received information on pregnancy care from their friends. These narratives showed that the emotional connection that they have formed with friends who share similar experiences of pregnancy has helped them respond directly to their emotional needs, which has contributed greatly to positive maternal care and wellbeing.

> “*My friends fell pregnant when we were still at school, and so we talk about everything. They are also open. I have open friends. So if we have challenges, we talk and help each other. So most of the information I get from them*.” (Participant 1, 21-25 years old).
>
> “*Some of the information I got it from my other friend. She has a baby so she has the experience*.” (Participant 18, 26-30 years old).

Some participants also received information support from their families around pregnancy care, through families imparting information on various traditional or cultural pregnancy practices that the participants had to follow. The women’s narratives around this showed that the cultural knowledge they possess around pregnancy has markedly influenced their pregnancy experiences and related meaning-making. In particular, it has resulted in young women accepting their pregnancies and protecting their health or pregnancies from a spiritual perspective.

> “*The cultural practices that we follow at home and in my church during pregnancy is that they tie you with a band on your waist to protect you, you drink water which they prayed over, in case you come across evil and being pregnant, the baby catches a lot of things*.” (Participant 1, 21-25 years old).
>
> “*My parent gave me remedies to drink and waist braces. My parent said they protect the child from evil spirits*.” (Participant 9, 16-20 years old).
>
> “*My family took me to ‘someone’ (traditional healer) just so they can ‘strengthen my pregnancy’. He said I must come with milk and eggs, ja and then he mixed them and he said I must bathe with them, and then he said I must also bring Vaseline as well, and then he mixed it I can’t remember what herbs he used, but then told me to lather my body with it every night*.” (Participant 13, 21-25 years old).
>
> “*There is a man that my parent took me to. He gave me water to drink. Water mixed with some soil and a string. It was to tie the pregnancy because they suspected that I might miscarry this one as well*.” (Participant 14, 21-25 years old)
>
> “*My elders do tell me that I don’t have to show my stomach off all the time because people out there can prevent me from delivering the baby and all that stuff; ja and also that I should wash myself with ‘isiwasho’ (traditional medicine infused water). It’s to protect the baby they say, ja and protect delivery*.” (Participant 16, 16-20 years old).

One participant reported that she received no information support from peers, health services, members of her family or from her friends, but gathered the information herself through her own learned experiences during her first pregnancy.

> “*For me it happened naturally. It was just general knowledge, having to understand things the way they should be. With my firstborn, I was breastfeeding him and I learned how to take care of him*.” (Participant 11, 21-25 years old).

## Discussion

The study’s objective was to explore the experiences of young females, in particular, receipt of social support, and its effect on pregnancy care and wellbeing during the prenatal period, through a phenomenological approach, in the context of Soweto, South Africa. The findings demonstrate that support from members of their various relationship networks contributed greatly to the young women’s pregnancy care and emotional wellbeing. Such support was found to be available to most participants of the study, even though the support that they received either from their partner or family, and the type of support that they received (instrumental, emotional and informational support) somewhat varied. This contributes greatly to young mothers having increased acceptance of their pregnancies and a greater sense of potential parental efficacy (36).

These findings address a critical gap in the literature, since the nature of social support has not previously been explored from a qualitative perspective in South Africa. It is pivotal to acquire a nuanced comprehension of these interactions and their role in maternal pregnancy care and wellbeing since many young women face uncertainty about their emotional and cognitive readiness in assuming a parental role (10). These cognitions and emotions do not exist in isolation but tend to be influenced by one’s interactions with various individuals namely friends, family members, sexual partners and community members (10). Previous studies in the South African context have mostly focused on the association between depressive symptomatology and social support in the prenatal period (30, 73, 74) and social support among pregnant young women, including women living with HIV (46, 75). Other quantitative research on social support in pregnancy has mostly been conducted in mainly in non-African high-income settings (10) where social support has been measured quantitatively and where measures of social support have been applied generically with lack of relevance to a particular population (76).

The novelty of this study was therefore its qualitative exploration of the type of social and its source perceived by young pregnant women in South Africa, and how this had an impact on their pregnancy care and wellbeing. Exploring pregnant young women’s social receipt within the context of Soweto given that the area is largely a historically underprivileged high-density periurban area in South Africa was vital, where young females are particularly afflicted by gender inequality, impoverishment, low educational attainment, poor or lack of social capital, and and economic disadvantages (77, 78). These social issues ultimately may expose them to gender-based violence and early pregnancy, which have both become the norm in South Africa (77, 78). Therefore, it was important to ground the narrative accounts of the young women within the specific sociocultural context of Soweto, and within their experiences. The application of an interpretive phenomenological approach has helped provide new insight as it has shed light on realities and meaning-making around pregnancy and receipt of social support in this context.

It was further found in the study that partner and family behavioural responses to pregnancy disclosure and type of social support that the young women received from these social networks, played a significant role in their emotional wellbeing as well as in the young women taking an initiative to focus on caring for their pregnancy and initiating antenatal care. This aligns with evidence indicating the major influence that sexual partners and family members have in young women’s pregnancy care, and their physical and psychological wellbeing (37, 79-85). Also, paternal or family support and stable relationships may mediate or combat distress on expectant women which may reduce the risk of experiencing a poor birth outcome (86). This type of support thus has the potential to increase their likelihood of experiencing positive birth outcomes and also encourages young women to engage in positive pregnancy care behaviours.

Our findings on father support are congruent to other literature which has shown that partner support, stability and dependability plays a key role in positive maternal behaviours and acceptance of pregnancy (87). Moreover, an engaged partner who cares about the process of pregnancy contributes positively to the psychological health and health behaviours of expectant females, which subsequently results in positive pregnancy experiences (88). Thus it can be suggested that young expectant females value the attachment that they have with their partners as they largely deem these supportive networks as playing a key role in helping them to cope better with their pregnancies. An unanticipated finding emerging from our findings was that most expectant females reported receiving emotional and instrumental support from their sexual partners following disclosure of their pregnancy, with partners accepting the pregnancies and having strengthened and stable relationships with their partners.

Male partners were mostly cited as the main sources of social support with the type of support received being of an emotional and instrumental nature. These findings seem contradictory to what has been observed in previous literature, which indicates that unmarried fathers have increased odds of being less involved during pregnancy than fathers who are in a marital union, thus having no obligation towards providing social support to the mother during pregnancy (89, 90). Our findings around partner support were also highly unanticipated given that intimate partner violence is another social ill in South Africa which has partly contributed to young women being exposed to early childbearing due to unequal power relations and the power of sexual coercion. This compromises women’s sexual health as women who are in relationships characterized by intimate partner violence cannot employ their autonomy when it comes to the prevention of pregnancy and contraceptive or condom use (90).

Expectant fathers have received limited attention in South African literature with respect to their supportive role during pregnancy (91-93) with studies viewing fatherhood as only starting postpartum (94, 95). Furthermore, South Africa is largely deemed as a country globally that is characterized by high levels of father absenteeism (96-98) which can be partly traced back to the apartheid regime, urbanization and labor migration, which had a strong impact on family life and composition and changed the manner in which families functioned, with these changes being more prominent in the roles of fathers, mothers and extended family members (99, 100). This lack of participation in parenthood stems from the fact that most young fathers never knew their fathers which has contributed greatly to their lack of guidance and experience on father roles and responsibilities (100). Other constructs of fatherhood deem male emotional and nurturing forms of support as “soft” behaviours (101) which suggests that fathers have to be emotionally distant (102). In addition, non-involvement of young fathers can be attributed to the increase in poorly paid, unprotected and insecure jobs and enduring rates of impoverishment and inequity in South Africa that place men at a disadvantage of being able to provide adequate care to their families (103, 104). Of note, a previous study focusing on out of work and casual working young males in an urban area of South Africa showed that some men have abandoned their paternal responsibilities on the grounds that child support grants in the country have come to substitute men’s roles as “providers” (105). Our findings regarding the role of partners’ support during pregnancy indicate that further research is warranted to explore this role further.

Although some participants in this study reported receiving emotional and instrumental support from their families, other participants reporting strained relationships with their families. A possible explanation for this is that families punish young girls who fall pregnant through giving them poor or no support which leads to challenges for young mothers to accept their pregnancies and engage in positive maternal behaviours (106). This is because premarital childbearing has been largely associated with social stigma and familial shame (107). Moreover, early childbearing among young women in particular, from a socioeconomic standpoint, has been found to have unfavorable results on the child, the young mother and extended family members (107-109). This is because when a young woman is unmarried, the young mother and her family shoulder the financial burden of childrearing (110).

The composition of household members and the number of members in the household have been found to affect fertility behaviour significantly (111). South Africa has dynamic household living arrangements, fewer people entering marital unions, and a rise in households in which adult females are the sole or main income generators and decision-makers (112). In South Africa, the role of the family has changed over time (100, 113) which has resulted in extensive changes in living arrangements and familial responsibilities (114) and some families experiencing major challenges in providing comprehensive care to their children, attributable to societal problems associated with poverty, large family sizes, the HIV/AIDS pandemic and dismantling of the nuclear family unit. These considerations need to be taken into account especially since these factors can all serve as a set of varied and inter-related factors that can influence fertility behaviour and outcomes of young women.

This current study found that other young women received support from both their sexual partners and families while other participants either reported receiving support only from their partners or only from their families. These findings suggest experiences of receipt of social support among young women differ greatly, with the type and source of social support not being universal. For instance, some young women experience pressure from their peers and forces to engage in sexual activities in fear of their partners who threaten to end the relationship often resulting in unintended pregnancies or termination (108, 115-117) while other young expectant females’ experience of pregnancy involves a denial of paternity (106). Conversely, other women have positive experiences due to their partners accepting paternity and having an increased need to have a relationship with their children, which is often a decision that is influenced by the partner’s family (67).

These experiences in turn affect young women’s acceptance or non-acceptance of their pregnancies given that the experience of childbearing among young women, its causes and consequences are not uniform (106). Given that early childbearing among young women is a phenomenon that is embedded within society (67) any changes that occur in society and any perceptions that members of communities hold about early childbearing among young women, affect how young expectant females experience pregnancy and motherhood (106). Also, complexities around communication pertaining to young women’s reproductive health increase their susceptibility to early childbearing and affect how they experience pregnancy. It is thus pivotal to consider the significance of each source of support and its well-being individually because pregnant young women are not a homogenous group. They have a variety of demographic and familial characteristics, different relations with their families and partners and the experiences and context in which they live may differ which contributes greatly to engaging in positive maternal behaviours or lack thereof.

Since experiences of pregnancy are vastly different and complexities around family and partner dynamics create different realities for these young women, social support needs to be relevant to the different realities of young women. Focusing attention on pregnant women to identify those who experience poor levels of social support along with the provision of community-based support services in collaboration with partners and families, may help foster positive behavioural outcomes with pregnant women taking an initiative to focus on caring for their pregnancy and initiating antenatal care. Furthermore, interventions that urge young women to use personal networks are needed. Health professionals can engage directly with partners and families of young women, by providing information and training on how to support the young women whilst being sensitive to the beliefs and cultural values of these sources of support. Health workers should also be encouraged to mobilize the broader communities of these young women through mass-media campaigns and discourses that promote the need for the supportive role of partners and families and also empower young women to have agency and autonomy over the decisions they make on their reproductive health.

The strength of this study is the application of the interpretive phenomenological approach, as it helped the researchers to gain a nuanced understanding of participants’ views and meaning-making around the phenomenon under study. This provides in-depth insights into a particular context, although transferability to other contexts should not be assumed, particularly since participants were part of a trial. The study’s limitation was that the support networks of the young women were not included as study participants. These individuals could also provide their perspectives on how they believe their provision of social support has had a major influence in the pregnancy care and wellbeing of the young mothers.

## Conclusion

The existence of social support has a major influence on pregnancy care and wellbeing and a lack thereof can pose significant risks on both maternal and child health. Focusing on social support could help contribute to reducing maternal mortality rates and achieving reproductive health accessibility universally. Pregnant women who receive sufficient social support from immediate networks have increased potential to embrace and give attention to pregnancy-related changes. This will, in turn, foster positive behavioural outcomes that encourage engaging in good pregnancy care practices and acceptance of motherhood. The important role of maternal support during pregnancy suggests that the wider community needs to be educated by policymakers and healthcare providers about the importance of partner, family and peer support in order to minimize risks that may affect maternal pregnancy care and wellbeing. Future research should be conducted that explores the dynamics of social support within various family structures. Improving receipt of social support by young mothers could enhance the promotion of physical and mental health of mothers which would subsequently result in the engagement of healthy behaviours in the perinatal period and positive birth outcomes.

## Data Availability

All data supporting our work is provided within the manuscript.

## Acknowledgements

This study was supported by the South African Medical Research Council, and the Canadian Institutes of Health Research. SAN is supported by the DSI-NRF Centre of Excellence in Human Development.

## Supporting information

**S1 Table. Themes focusing on young women’s experiences of social support on pregnancy care and wellbeing**.

**S2 Table. Demographic Profile of the participants.**

**S3 Interview guide**.

## Data Availability Statement

All data supporting our work is provided within the manuscript

## Funding

This study was supported by the South African Medical Research Council and DSI-NRF Centre of Excellence in Human Development at the University of the Witwatersrand, Johannesburg. The funders had no role in preparation of the manuscript, study design, data collection and analysis.

## Competing interests

The authors declare that they have no competing interests

## Notes

### Competing Interest Statement

The authors have declared no competing interest.

### Author Declarations

Approval to conduct the study was granted by the Human Research Ethics Committee (Medical) based at the University of the Witwatersrand (M190449). Respondents gave written permission to be interviewed, and for the narratives to be recorded.

## References

1. Agostini F, Neri E, Salvatori P, Dellabartola S, Bozicevic L, Monti F. Antenatal depressive symptoms associated with specific life events and sources of social support among Italian women. Maternal and child health journal. 2015;19(5):1131–41.

2. Skurzak A, Kicia M, Wiktor K, Iwanowicz-Palus G, Wiktor H. Social support for pregnant women. Pol J Public Health. 2015;125:169–72.

3. Zauszniewski JA. Stress experiences and mental health of pregnant women: the mediating role of social support. Issues in mental health nursing. 2019.

4. McLeish J, Redshaw M. Mothers’ accounts of the impact on emotional wellbeing of organised peer support in pregnancy and early parenthood: a qualitative study. BMC pregnancy and childbirth. 2017;17(1):1–14.

5. Diego MA, Field T, Hernandez-Reif M, Cullen C, Schanberg S, Kuhn C. Prepartum, postpartum, and chronic depression effects on newborns. Psychiatry: Interpersonal and Biological Processes. 2004;67(1):63–80.

6. Chang S-R, Kenney NJ, Chao Y-MY. Transformation in self-identity amongst Taiwanese women in late pregnancy: a qualitative study. International Journal of Nursing Studies. 2010;47(1):60–6.

7. Ibanez G, Blondel B, Prunet C, Kaminski M, Saurel-Cubizolles M-J. Prevalence and characteristics of women reporting poor mental health during pregnancy: findings from the 2010 French National Perinatal Survey. Revue d’Épidémiologie et de Santé Publique. 2015;63(2):85–95.

8. Lagadec N, Steinecker M, Kapassi A, Magnier AM, Chastang J, Robert S, et al. Factors influencing the quality of life of pregnant women: a systematic review. BMC pregnancy and childbirth. 2018;18(1):114.

9. Alzboon G, Vural G. The Experience of Healthy Pregnancy in High Parity Women: A Phenomenological Study in North Jordan. Medicina. 2021;57(8):853.

10. Mbatha K. Social support as psychological mediator among African black women who have recently given birth: Citeseer; 2014.

11. Rini C, Schetter CD, Hobel CJ, Glynn LM, Sandman CA. Effective social support: Antecedents and consequences of partner support during pregnancy. Personal Relationships. 2006;13(2):207–29.

12. Reblin M, Uchino BN. Social and emotional support and its implication for health. Current opinion in psychiatry. 2008;21(2):201.

13. Edmonds JK, Paul M, Sibley LM. Type, content, and source of social support perceived by women during pregnancy: Evidence from Matlab, Bangladesh. Journal of health, population, and nutrition. 2011;29(2):163.

14. Abadi MNL. Social support, Coping, and Self-Esteem in Relation to psychosocial factors: a study of health issues and birth weight in young mothers in Tehran, Iran: Department of Social Work, Umeå University; 2012.

15. Maharlouei N. The importance of social support during pregnancy. Women’s Health Bulletin. 2016;3(1):1–.

16. Milgrom J, Hirshler Y, Reece J, Holt C, Gemmill AW. Social support—a protective factor for depressed perinatal women? International journal of environmental research and public health. 2019;16(8):1426.

17. Rafiei N, Amini Rarani M, Eizadi F, Rafiey H, Seyedghasemi NS. Social support and its role in the prevention of depression and anxiety during pregnancy in Turkmen women. International Journal of Biomedicine and Public Health. 2019;2(4):75–80.

18. Asselmann E, Kunas SL, Wittchen H-U, Martini J. Maternal personality, social support, and changes in depressive, anxiety, and stress symptoms during pregnancy and after delivery: A prospectivelongitudinal study. Plos one. 2020;15(8):e0237609.

19. Nakamura A, Sutter-Dallay A-L, El-Khoury Lesueur F, Thierry X, Gressier F, Melchior M, et al. Informal and formal social support during pregnancy and joint maternal and paternal postnatal depression: data from the French representative ELFE cohort study. International Journal of Social Psychiatry. 2020;66(5):431–41.

20. Mountain RV, Zhu Y, Pickett OR, Lussier AA, Goldstein JM, Roffman JL, et al. Association of Maternal Stress and Social Support During Pregnancy With Growth Marks in Children’s Primary Tooth Enamel. JAMA network open. 2021;4(11):e2129129–e.

21. Renbarger KM, Place JM, Schreiner M. The Influence of Four Constructs of Social Support on Pregnancy Experiences in Group Prenatal Care. Women’s Health Reports. 2021;2(1):154–62.

22. Robertson E, Grace S, Wallington T, Stewart DE. Antenatal risk factors for postpartum depression: a synthesis of recent literature. General hospital psychiatry. 2004;26(4):289–95.

23. Figueiredo B, Bifulco A, Pacheco A, Costa R, Magarinho R. Teenage pregnancy, attachment style, and depression: A comparison of teenage and adult pregnant women in a Portuguese series. Attachment & Human Development. 2006;8(2):123–38.

24. Lee AM, Lam SK, Lau SMSM, Chong CSY, Chui HW, Fong DYT. Prevalence, course, and risk factors for antenatal anxiety and depression. Obstetrics & Gynecology. 2007;110(5):1102–12.

25. Littleton HL, Breitkopf CR, Berenson AB. Correlates of anxiety symptoms during pregnancy and association with perinatal outcomes: a meta-analysis. American journal of obstetrics and gynecology. 2007;196(5):424–32.

26. Leigh B, Milgrom J. Risk factors for antenatal depression, postnatal depression and parenting stress. BMC psychiatry. 2008;8(1):1–11.

27. Wahn EH, Nissen E. Sociodemographic background, lifestyle and psychosocial conditions of Swedish teenage mothers and their perception of health and social support during pregnancy and childbirth. Scandinavian journal of public health. 2008;36(4):415–23.

28. Grote NK, Bridge JA, Gavin AR, Melville JL, Iyengar S, Katon WJ. A meta-analysis of depression during pregnancy and the risk of preterm birth, low birth weight, and intrauterine growth restriction. Archives of general psychiatry. 2010;67(10):1012–24.

29. Rashid A, Mohd R. Poor social support as a risk factor for antenatal depressive symptoms among women attending public antennal clinics in Penang, Malaysia. Reproductive Health. 2017;14(1):1–8.

30. Bedaso A, Adams J, Peng W, Sibbritt D. The relationship between social support and mental health problems during pregnancy: a systematic review and meta-analysis. Reproductive health.2021;18(1):1–23.

31. Kim TH, Connolly JA, Tamim H. The effect of social support around pregnancy on postpartum depression among Canadian teen mothers and adult mothers in the maternity experiences survey. BMC pregnancy and childbirth. 2014;14(1):1–9.

32. Maidaliza A, Susanti SS. Social Support Received by Postpartum Mothers in Indonesia: A Descriptive Phenomenological Study. 2020.

33. Mbekenga CK, Lugina HI, Christensson K, Olsson P. Postpartum experiences of first-time fathers in a Tanzanian suburb: a qualitative interview study. Midwifery. 2011;27(2):174–80.

34. Shefer T, Bhana D, Morrell R. Teenage pregnancy and parenting at school in contemporary South African contexts: Deconstructing school narratives and understanding policy implementation. Perspectives in Education. 2013;31(1):1–10.

35. VanDenBerg MP. Protective factors for teen mothers: relations among social support, psychological resources, and child rearing practices: Colorado State University; 2012.

36. Coert SL, Adebiyi BO, Rich E, Roman NV. A comparison of the relationship between parental efficacy and social support systems of single teen mothers across different family forms in South African low socioeconomic communities. BMC women’s health. 2021;21(1):1–11.

37. Leahy-Warren P, McCarthy G, Corcoran P. First-time mothers: social support, maternal parental self-efficacy and postnatal depression. Journal of clinical nursing. 2012;21(3-4):388–97.

38. Morikawa M, Okada T, Ando M, Aleksic B, Kunimoto S, Nakamura Y, et al. Relationship between social support during pregnancy and postpartum depressive state: a prospective cohort study. Scientific reports. 2015;5(1):1–9.

39. Ginja S, Coad J, Bailey E, Kendall S, Goodenough T, Nightingale S, et al. Associations between social support, mental wellbeing, self-efficacy and technology use in first-time antenatal women: data from the BaBBLeS cohort study. BMC pregnancy and childbirth. 2018;18(1):1–11.

40. Ngui E, Cortright A, Blair K. An investigation of paternity status and other factors associated with racial and ethnic disparities in birth outcomes in Milwaukee, Wisconsin. Maternal and Child Health Journal. 2009;13(4):467–78.

41. Alio AP, Kornosky JL, Mbah AK, Marty PJ, Salihu HM. The impact of paternal involvement on feto-infant morbidity among Whites, Blacks and Hispanics. Maternal and child health journal. 2010;14(5):735–41.

42. Alio AP, Mbah AK, Kornosky JL, Wathington D, Marty PJ, Salihu HM. Assessing the impact of paternal involvement on racial/ethnic disparities in infant mortality rates. Journal of community health. 2011;36(1):63–8.

43. Ilska M, Przybyła-Basista H. Partner support as a mediator of the relationship between prenatal concers and psychological well-being in pregnant women. 2017.

44. Falade-Fatila O, Adebayo AM. Male partners’ involvement in pregnancy related care among married men in Ibadan, Nigeria. Reproductive health. 2020;17(1):1–12.

45. Kumar SA, Brock RL, DiLillo D. Partner support and connection protect couples during pregnancy: A daily diary investigation. Journal of Marriage and Family. 2021.

46. Hill LM, Maman S, Groves AK, Moodley D. Social support among HIV-positive and HIV-negative adolescents in Umlazi, South Africa: changes in family and partner relationships during pregnancy and the postpartum period. BMC Pregnancy and childbirth. 2015;15(1):1–9.

47. Granville G, Sugarman W. Someone in my corner: a volunteer peer support programme for pregnancy, birth and beyond. Final Evaluation Report In Parents 1st. 2012.

48. Montgomery P, Mossey S, Adams S, Bailey PH. Stories of women involved in a postpartum depression peer support group. International Journal of Mental Health Nursing. 2012;21(6):524–32.

49. Liu M-C, Kuo S-H, Lin C-P, Yang Y-M, Chou F-H, Yang Y-H. Effects of professional support on nausea, vomiting, and quality of life during early pregnancy. Biological research for nursing. 2014;16(4):378–86.

50. Ekström AC, Thorstensson S. Nurses and midwives professional support increases with improved attitudes-design and effects of a longitudinal randomized controlled process-oriented intervention. BMC pregnancy and childbirth. 2015;15(1):1–9.

51. Bäckström C. Professional and social support for first-time mothers and partners during childbearing: Jönköping University, School of Health and Welfare; 2018.

52. Akinwaare MO, Ogbeye GB, Ejimofor N. Social Support During Pregnancy among Pregnant Women in Ibadan, Nigeria. International Journal of Nursing, Midwife and Health Related Cases. 2019;5(1):14–26.

53. Hijazi HH, Alyahya MS, Al Abdi RM, Alolayyan MN, Sindiani AM, Raffee LA, et al. The Impact of Perceived Social Support During Pregnancy on Postpartum Infant-Focused Anxieties: A Prospective Cohort Study of Mothers in Northern Jordan. International journal of women’s health. 2021;13:973.

54. Nicholls D. Qualitative research: part one–philosophies. International Journal of Therapy and Rehabilitation. 2009;16(10):526–33.

55. Tuohy D, Cooney A, Dowling M, Murphy K, Sixsmith J. An overview of interpretive phenomenology as a research methodology. Nurse researcher. 2013;20(6).

56. Abota TL, Gashe FE, Kabeta ND. Postpartum Women’s Lived Experiences of Perinatal Intimate Partner Violence in Wolaita Zone, Southern Ethiopia: A Phenomenological Study Approach. International journal of women’s health. 2021;13:1103.

57. Flood A. Understanding phenomenology. Nurse researcher. 2010;17(2).

58. Maggs-Rapport F. Combining methodological approaches in research: ethnography and interpretive phenomenology. Journal of advanced nursing. 2000;31(1):219–25.

59. Arghavanian FE, Roudsari RL, Heydari A, Bahmani MND. Men’s confrontation with pregnancy from women’s point of view: an ethno phenomenological approach. Journal of Caring Sciences. 2019;8(4):231.

60. Lundgren I, Wahlberg V. The experience of pregnancy: a hermeneutical/phenomenological study. The Journal of Perinatal Education. 1999;8(3):12–20.

61. Giorgi A. One type of analysis of descriptive data: Procedures involved in following a scientific phenomenological method. Methods. 1989;1(3):39–61.

62. Groenewald T. A phenomenological research design illustrated. International journal of qualitative methods. 2004;3(1):42–55.

63. Draper C, Prioreschi A, Ware L, Lye S, Norris S. Pilot implementation of Bukhali: A preconception health trial in South Africa. SAGE Open Medicine. 2020;8:2050312120940542.

64. Grønlund J. The genesis of a football field: urban football in Soweto, South Africa. Soccer & Society. 2021;22(3):218–30.

65. Battersby J, McLachlan M. Urban food insecurity: A neglected public health challenge. South African Medical Journal. 2013;103(10):716–7.

66. Miller CL, Nkala B, Closson K, Chia J, Cui Z, Palmer A, et al. The Botsha Bophelo adolescent health study: a profile of adolescents in Soweto, South Africa. Southern African journal of HIV medicine. 2017;18(1):1–10.

67. Jewkes R, Morrell R, Christofides N. Empowering teenagers to prevent pregnancy: lessons from South Africa. Culture, health & sexuality. 2009;11(7):675–88.

68. Taylor M, Jinabhai C, Dlamini S, Sathiparsad R, Eggers MS, De Vries H. Effects of a teenage pregnancy prevention program in KwaZulu-Natal, South Africa. Health care for women international. 2014;35(7-9):845–58.

69. Amod Z, Halana V, Smith N. School-going teenage mothers in an under-resourced community: lived experiences and perceptions of support. Journal of Youth Studies. 2019;22(9):1255–71.

70. Software V. MAXQDA 2020. VERBI Software Berlin; 2019.

71. Wirihana L, Welch A, Williamson M, Christensen M, Bakon S, Craft J. Using Colaizzi’s method of data analysis to explore the experiences of nurse academics teaching on satellite campuses. Nurse Researcher (2014+). 2018;25(4):30.

72. Kr P. Application of Colaizzi’s Method of Data Analysis in Phenomenological Research. MedicoLegal Update. 2021;21(2).

73. Brittain K, Mellins CA, Phillips T, Zerbe A, Abrams EJ, Myer L, et al. Social support, stigma and antenatal depression among HIV-infected pregnant women in South Africa. AIDS and Behavior. 2017;21(1):274–82.

74. Smith M, Mitchell AS, Townsend ML, Herbert JS. The relationship between digital media use during pregnancy, maternal psychological wellbeing, and maternal-fetal attachment. PloS one. 2020;15(12):e0243898.

75. Mfusi S, Mahabeer M. Psychosocial adjustment of pregnant women infected with HIV/AIDS in South Africa. Journal of Psychology in Africa. 2000;10(2):122–45.

76. McDowell I. Measuring health: a guide to rating scales and questionnaires: Oxford University Press, USA; 2006.

77. Jewkes R, Vundule C, Maforah F, Jordaan E. Relationship dynamics and teenage pregnancy in South Africa. Social science & medicine. 2001;52(5):733–44.

78. Bhana D. “Girls are not free”—In and out of the South African school. International Journal of Educational Development. 2012;32(2):352–8.

79. Turner RJ, Sorenson AM, Turner JB. Social contingencies in mental health: A seven-year followup study of teenage mothers. Journal of Marriage and Family. 2000;62(3):777–91.

80. Serovich JM, Kimberly J, Mosack K, Lewis T. The role of family and friend social support in reducing emotional distress among HIV-positive women. AIDS care. 2001;13(3):335–41.

81. Glazier R, Elgar F, Goel V, Holzapfel S. Stress, social support, and emotional distress in a community sample of pregnant women. Journal of Psychosomatic Obstetrics & Gynecology. 2004;25(34):247–55.

82. Milan S, Ickovics JR, Kershaw T, Lewis J, Meade C, Ethier K. Prevalence, course, and predictors of emotional distress in pregnant and parenting adolescents. Journal of Consulting and Clinical Psychology. 2004;72(2):328.

83. Rahman A, Chowdhury S. Determinants of chronic malnutrition among preschool children in Bangladesh. Journal of biosocial science. 2007;39(2):161–73.

84. Kathree T, Petersen I. South African Indian women screened for postpartum depression: a multiple case study of postpartum experiences. South African Journal of Psychology. 2012;42(1):37–50.

85. Stapleton LRT, Schetter CD, Westling E, Rini C, Glynn LM, Hobel CJ, et al. Perceived partner support in pregnancy predicts lower maternal and infant distress. Journal of family psychology. 2012;26(3):453.

86. Shah R, Mullany LC, Darmstadt GL, Mannan I, Rahman SM, Talukder RR, et al. Incidence and risk factors of preterm birth in a rural Bangladeshi cohort. BMC pediatrics. 2014;14(1):1–11.

87. Calderwood L, Kiernan K, Joshi H, Smith K, Ward K, Dex S, et al. Parenthood and parenting. Children of the 21st century: from birth to nine months. 2005.

88. Alio AP, Lewis CA, Scarborough K, Harris K, Fiscella K. A community perspective on the role of fathers during pregnancy: a qualitative study. BMC pregnancy and childbirth. 2013;13(1):1–11.

89. Cabrera N, Brooks-Gunn J, Moore K, West J, Boller K, Tamis-LeMonda CS. Bridging research and policy: Including fathers of young children in national studies. Handbook of father involvement: Routledge; 2012. p. 492–526.

90. Bottoman PE. Pregnant women’s construction of social support from their intimate partners during pregnancy: Rhodes University; 2018.

91. Richter L. The importance of fathering for children. Baba: men and fatherhood in South Africa. 2006:53–69.

92. Maman S, Moodley D, Groves AK. Defining male support during and after pregnancy from the perspective of HIV-Positive and HIV-Negative women in Durban, South Africa. Journal of midwifery & women’s health. 2011;56(4):325–31.

93. Groves AK, McNaughton-Reyes HL, Foshee VA, Moodley D, Maman S. Relationship factors and trajectories of intimate partner violence among South African women during pregnancy and the postpartum period. PLoS One. 2014;9(9):e106829.

94. Clowes L. Men and children: changing constructions of fatherhood in Drum magazine, 19511965. HSRC Press; 2006.

95. Ratele K, Shefer T, Clowes L. Talking South African fathers: A critical examination of men’s constructions and experiences of fatherhood and fatherlessness. South African journal of psychology. 2012;42(4):553–63.

96. Richter L, Chikovore J, Makusha T. The status of fatherhood and fathering in South Africa. Childhood education. 2010;86(6):360–5.

97. Richter L, Desmond C, Hosegood V, Madhavan S, Makiwane M, Makusha T, et al. Fathers and other men in the lives of children and families. 2012.

98. Freeks FE. Die noodsaak van Skrifgefundeerde vaderskap as antwoord op die voortslepende probleem van vaderafwesigheid in Suid-Afrika. Journal for Christian Scholarship= Tydskrif vir Christelike Wetenskap. 2016;52(1):1–27.

99. Smit R. The impact of labor migration on African families in South Africa: Yesterday and today. Journal of Comparative Family Studies. 2001;32(4):533–48.

100. Sooryamoorthy R, Makhoba M. The family in modern South Africa: Insights from recent research. Journal of Comparative Family Studies. 2016;47(3):309–21.

101. Gregory A, Milner S. What is “new” about fatherhood? The social construction of fatherhood in France and the UK. Men and masculinities. 2011;14(5):588–606.

102. Šmídová I. Fatherhood as a social construction: Mapping the challenges to promoting good fatherhood. European Fatherhood. 2007.

103. Barchiesi F. Wage labor, precarious employment, and social inclusion in the making of South Africa’s postapartheid transition. African Studies Review. 2008;51(2):119–42.

104. Barchiesi F. Precarious liberation: Workers, the state, and contested social citizenship in postapartheid South Africa: Suny Press; 2011.

105. Dawson HJ, Fouksman E. Labour, laziness and distribution: work imaginaries among the South African unemployed. Africa. 2020;90(2):229–51.

106. Mkhwanazi N. Revisiting the dynamics of early childbearing in South African townships. Culture, health & sexuality. 2014;16(9):1084–96.

107. Johnson-Hanks J. The lesser shame: abortion among educated women in southern Cameroon. Social science & medicine. 2002;55(8):1337–49.

108. Macleod CI, Tracey T. A decade later: follow-up review of South African research on the consequences of and contributory factors in teen-aged pregnancy. South African Journal of Psychology. 2010;40(1):18–31.

109. Ndinda C, Ndhlovu T, Khalema NE. Conceptions of contraceptive use in rural KwaZulu-Natal, South Africa: lessons for programming. International Journal of Environmental Research and Public Health. 2017;14(4):353.

110. Jewkes R, Christofides N. Teenage pregnancy: Rethinking prevention. HSRC Youth Policy Initiative Roundtable. 2008;5.

111. Mönkediek B, Bras H. Family systems and fertility intentions: Exploring the pathways of influence. European Journal of Population. 2018;34(1):33–57.

112. Hall K, Richter L, Mokomane Z, Lake L. South African Child Gauge 2018 children, families and the state collaboration and contestation. University of Cape Town: Children’s Institute.[Online]. Available at: http …; 2018.

113. Mokone J. Challenges experienced by grandparents raising grandchildren: An exploratory study. Social Work/Maatskaplike Werk. 2006;42(2).

114. Mathambo V, Gibbs A. Extended family childcare arrangements in a context of AIDS: collapse or adaptation? AIDS care. 2009;21(sup1):22–7.

115. Gevers A, Jewkes R, Mathews C, Flisher A. ‘I think it’s about experiencing, like, life’: a qualitative exploration of contemporary adolescent intimate relationships in South Africa. Culture, Health & Sexuality. 2012;14(10):1125–37.

116. Morrell R, Bhana D, Shefer T. Books and babies: Pregnancy and young parents in schools: HSRC Press Cape Town; 2012.

117. Miller E, Decker MR, McCauley HL, Tancredi DJ, Levenson RR, Waldman J, et al. Pregnancy coercion, intimate partner violence and unintended pregnancy. Contraception. 2010;81(4):31622.

